# Optimizing Protocol Efficiency in F-18 Flurpiridaz PET MPI Through Dose Ratio–Driven Reduction of Residual Activity

**DOI:** 10.1101/2025.09.02.25334885

**Authors:** Maria Alwan, Ahmad El Yaman, Mahmoud Al Rifai, Sandra Escobar, Shah F. Abbasi, Mohamad G. Ghosn, Marcelo F. Di Carli, Mouaz Al-Mallah

## Abstract

**Background:** F-18 Flurpiridaz is a newly FDA-approved tracer for PET MPI with a long half-life and improved physical characteristics. However, its long half-life leads to residual rest activity. Prior trials used 30 to 60 minutes delays between rest and stress injections and proposed stress-to-rest dose ratios of 1:2 to 1:3 to mitigate the potential impact of residual rest counts on the stress myocardial perfusion images.

**Objective:** To evaluate the effect of stress-to-rest dose ratio and time interval between injections on residual activity.

**Methods:** We analyzed consecutive 115 patients who underwent PET MPI with F-18 Flurpiridaz. Relative Residual activity was calculated as the ratio of tissue activity concentration (kBq/mL) in the first stress frame to that in the stress tissue phase. Linear regression was used to assess the association of dose ratio and injection time interval with global residual activity. The optimal dose ratio cutoff was identified. Mixed-effects models with interaction terms were used to assess whether the effect varied across vascular territories.

**Results:** A total of 115 patients underwent PET MPI. Results showed that increasing the stress-to-rest dose ratio was significantly associated with lower relative residual activity (β = -3.41; 95% CI: -4.72; -2.11 per 1-unit increase), while the time interval between injections showed no significant association (β =0.03 95% CI: -0.12; 0.17 per 1-minute increase). The optimal cutoff dose ratio to achieve relative residual activity <20% ranged between 3.2 and 4.3.

**Conclusion:** Increasing the stress-to-rest dose ratio between 3.2 and 4.3, effectively reduces residual activity to <20% of the stress dose across all time intervals, thereby enabling back-to-back imaging and improving protocol efficiency.

## Introduction

F-18 Flurpiridaz is a newly FDA-approved radiotracer for positron emission tomography (PET) myocardial perfusion imaging (MPI) with favorable physical-relatively long half-life and a short positron range- and pharmacokinetic characteristics featuring a high myocardial extraction fraction.(1-3) However, its imaging protocol is longer than that of traditional Rubidium-82 PET imaging, primarily due to the recommended 30-60-minute delay between the rest and stress injections for pharmacologic- and exercise-stress, respectively.(4, 5) The proposed delay between the rest and stress injections aim to reduce the impact of residual activity from the rest dose, preventing it from “shining through” into the stress images and thereby preserving optimal image contrast and interpretation. While widely adopted, these recommended delays are based on empirical data and prolong the imaging protocol, reducing workflow efficiency. Given the long half-life of F-18 of 110 minutes, we hypothesized that these standard wait times may be insufficient to meaningfully reduce residual counts. Therefore, we evaluated the impact of varying both the time interval between injections and the stress-to-rest dose ratio on residual activity, with the goal of optimizing Flurpiridaz PET protocols to enable back-to-back rest and stress imaging.

## Methods

### Study Population

The study population consisted of 115 consecutive patients who underwent a clinically indicated PET myocardial perfusion imaging (MPI) between February and August 2025 at Houston Methodist DeBakey Heart and Vascular Center. No patients were excluded from this cohort. Institutional Review Board (IRB) approval was obtained from the Houston Methodist Academic Institute prior to study initiation.

### Flurpiridaz Myocardial Perfusion PET/CT Imaging Protocol

Patients were scanned using a Biograph Vision PET/CT scanner (Siemens Healthineers, Knoxville, TN, USA). Patients were instructed to fast for 6 hours and to abstain from coffee and other caffeinated beverages for 18 hours prior to imaging. PET images were acquired in 3D list mode with 10 to 15 minutes of acquisition at rest and 10 to 15 minutes following pharmacologic stress. Low-dose, non-gated computed tomography (CT) scans were obtained for attenuation correction during both rest and stress imaging.

F-18 Flurpiridaz (F-18) was used exclusively as the myocardial perfusion radiotracer. F-18 was administered intravenously at rest and stress using variable doses and time intervals between the two injections to test different combinations of stress-to-rest dose ratios and waiting intervals between the two injections. Images were reconstructed and processed for static, gated, and dynamic imaging using standard clinical protocols. For the stress acquisition, the PET scanner was started before the intravenous injection of F-18 to capture the residual counts from the rest dose. Vasodilator stress was achieved with 0.4 mg of regadenoson (5 mL) administered intravenously over 10 seconds. Heart rate, blood pressure, and electrocardiogram (ECG) were monitored at baseline and at one-minute intervals throughout the stress test.

### Residual activity measurements

Residual tissue activity concentration (kBq/mL) from the rest dose was measured in the first frame of the stress dynamic series. Stress activity concentration was measured in the tissue phase globally, by coronary territory, and by segment using the standard 17-segment model.

Relative residual activity (RRA) was calculated as the ratio of tissue concentration in the first stress frame to that in the tissue phase and expressed as a percentage. A cutoff of ≤20% relative residual activity was used, as adequate reduction in residuals from the rest dose based on expert consensus. All measurements were performed using Corridor4DM software (Corridor4DM; Invia, Ann Arbor, Michigan).

### Variables of interest

The primary covariates of interest included the stress-to-rest dose ratio, the time interval between rest and stress injections, and the RRA. Additional covariates included sociodemographic characteristics (age, sex, and race) and clinical comorbidities (hypertension, dyslipidemia, and diabetes). Data on sociodemographic factors and cardiovascular-related conditions was collected by patient interview at the time of the test and supplemented by a review of the electronic health records by trained nurses.

### Statistical Analysis

Continuous variables were presented as median with interquartile range, and categorical variables were presented as the number with percentages. The association between the dose ratio and the time interval between rest and stress injections with RRA was evaluated using linear regression models. Further, the interaction between time interval and dose ratio was evaluated to determine whether the effect of one was dependent on the other. Predicted RRA was calculated across variant combinations of time interval and dose ratio. To identify optimal cutoff values for stress-to-rest dose ratio, we used logistic regression to determine the dose ratio at which the predicted probability of RRA <20% exceeded 90%, 95%, and 99%. Additionally, we evaluated cutoffs using linear regression using the 95% confidence interval (the range where the mean of the population falls) and the 95% prediction interval (the range within which an individual’s residual is likely to fall). These results are shown in the **supplement**. Further, we assessed the predicted RRA at varying time intervals while fixing the dose ratio at predefined thresholds (e.g.3.2). Finally, a mixed-effects model with a random intercept for each patient was fitted, incorporating an interaction term between dose ratio and vascular territory to evaluate whether the relationship between dose ratio and RRA varied across coronary territories.

All analyses were performed using R, version 4.4.1 (R Foundation for Statistical Computing, Vienna, Austria). The “lme4” package was used for mixed effect modeling and the “ggplot2” package for illustration. A p-value <0.05 was considered statistically significant.

## Results

A total of 115 patients were included, with a median (IQR) age of 74 years (68-78); 40 % were females and 77.4% were Caucasian. Among these, 84.3% had hypertension, 44.3% diabetes, and 95.7% dyslipidemia. The median (IQR) rest dose was 2.1 mCi (1.8–2.3), the stress dose was 5.5 mCi (5.2–5.8), and the stress to rest dose ratio was 2.6 (2.4–3.0). The median time interval between rest and stress injections was 19.4 minutes (17.1–21.4). The median global activity concentration were 9,724 kBq/mL (IQR, 7,819-11,963) and 54,166 kBq/mL (IQR, 44,452-65,579) in the first stress frame and the stress tissue phase, respectively, with a median global RRA of 18.2% (IQR, 15.1%-20.7%). Median RRA was 18.4% (IQR, 15.3-20.2) in the left anterior descending (LAD) territory; 17.9% (IQR, 15.2-20.9) in the left circumflex (LCx); and 17.9 % (IQR, 14.8-20.7) in the right coronary artery (RCA).

### Effect of time between doses and stress-to-rest dose ratio on relative residual activity

There was no association between a longer time interval between injections and RRA per 1-minute increase (β =0.03 95% CI: -0.12; 0.17, **Figure 1A**). In contrast, a higher stress-to-rest dose ratio was significantly associated with lower RRA per 1-unit increase (β = -3.41; 95% CI: - 4.72; -2.11, **Figure 1B**). When both variables were included in the same model, time interval between doses remained non-significant (β =-0.11; 95% CI: -0.25; 0.03), while the stress-to-rest dose ratio was significantly associated with lower RRA (β = -3.79; 95% CI: -5.17; -2.41). **Figure 2** displays a heatmap of predicted RRA based on the to stress-to-rest dose ratio and time interval. Vertically, the color gradient changes significantly, indicating that increasing the stress-to-rest dose ratio results in lower RRA for any given time interval. The interaction between dose ratio and time interval was not statistically significant (P = 0.75).

**Figure 1A.**
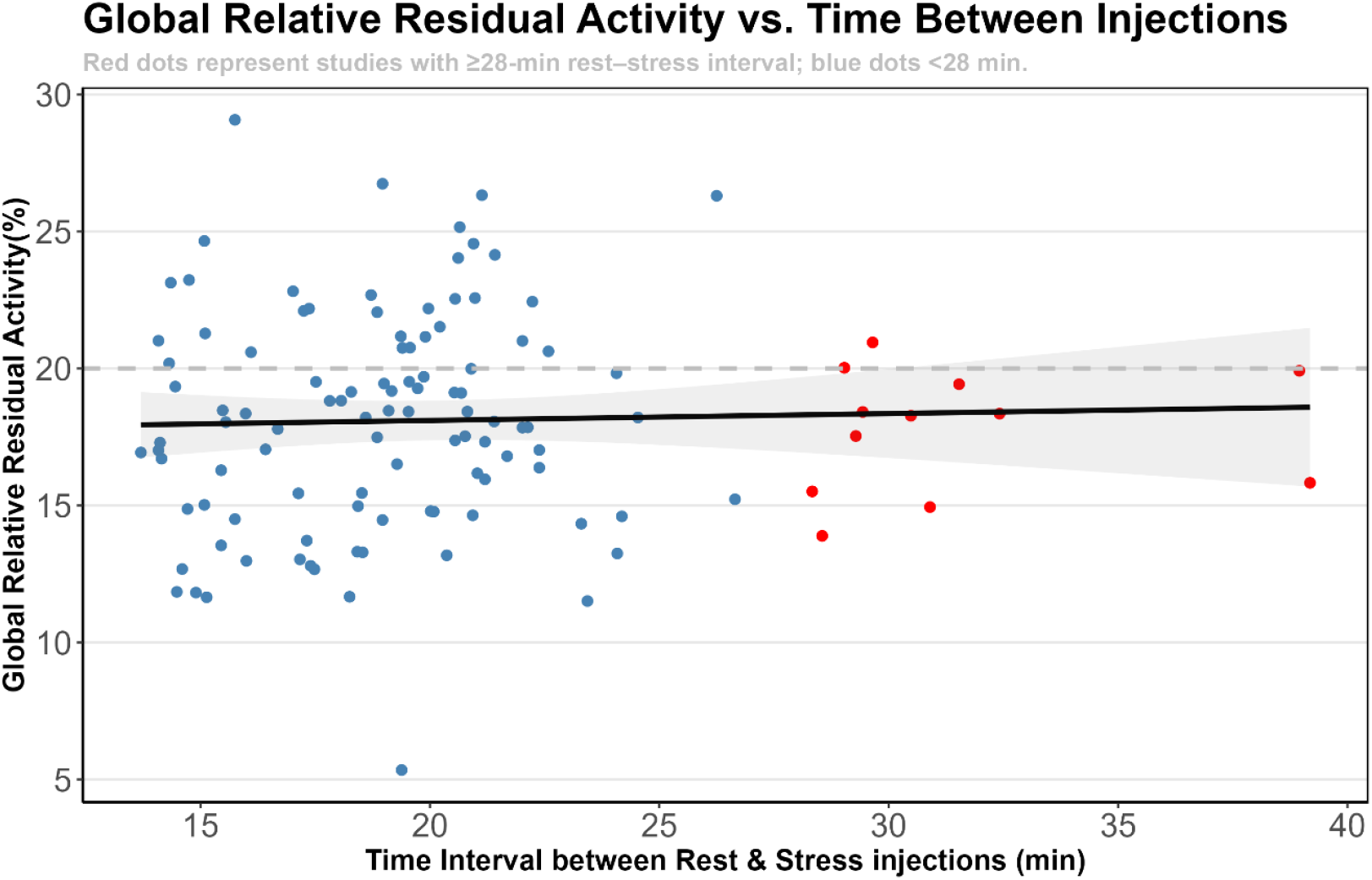
Association between time interval between rest and stress injections (minutes) and global relative residual activity (%). Red and blue dots represent studies with ≥28-minute and <28-minute delays between rest and stress injections, respectively.

**Figure 1B.**
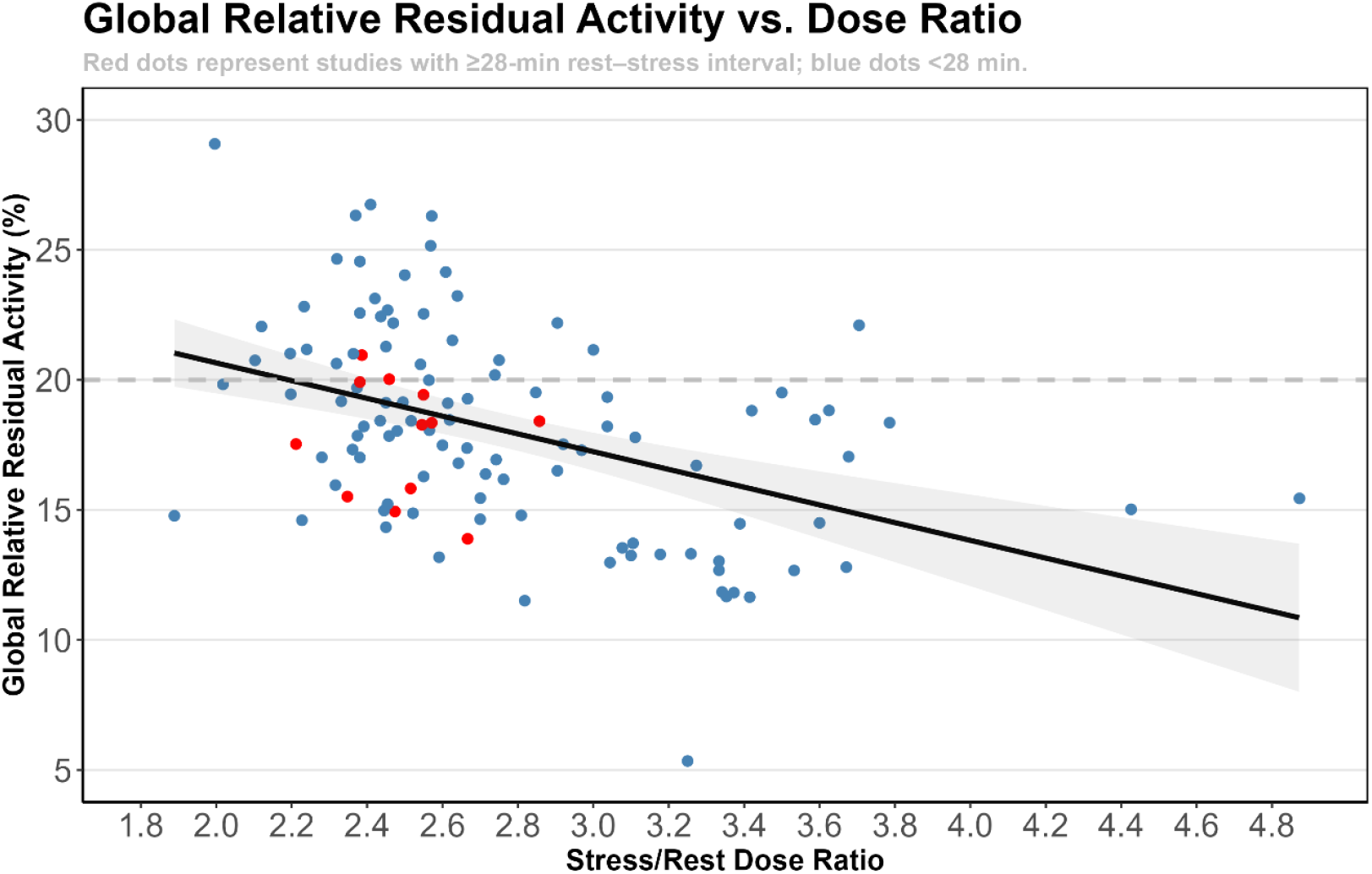
Association between stress-to-rest dose ratio and global relative residual activity (%). Red and blue dots represent studies with ≥28-minute and <28-minute delays between rest and stress injections, respectively.

**Figure 2.**
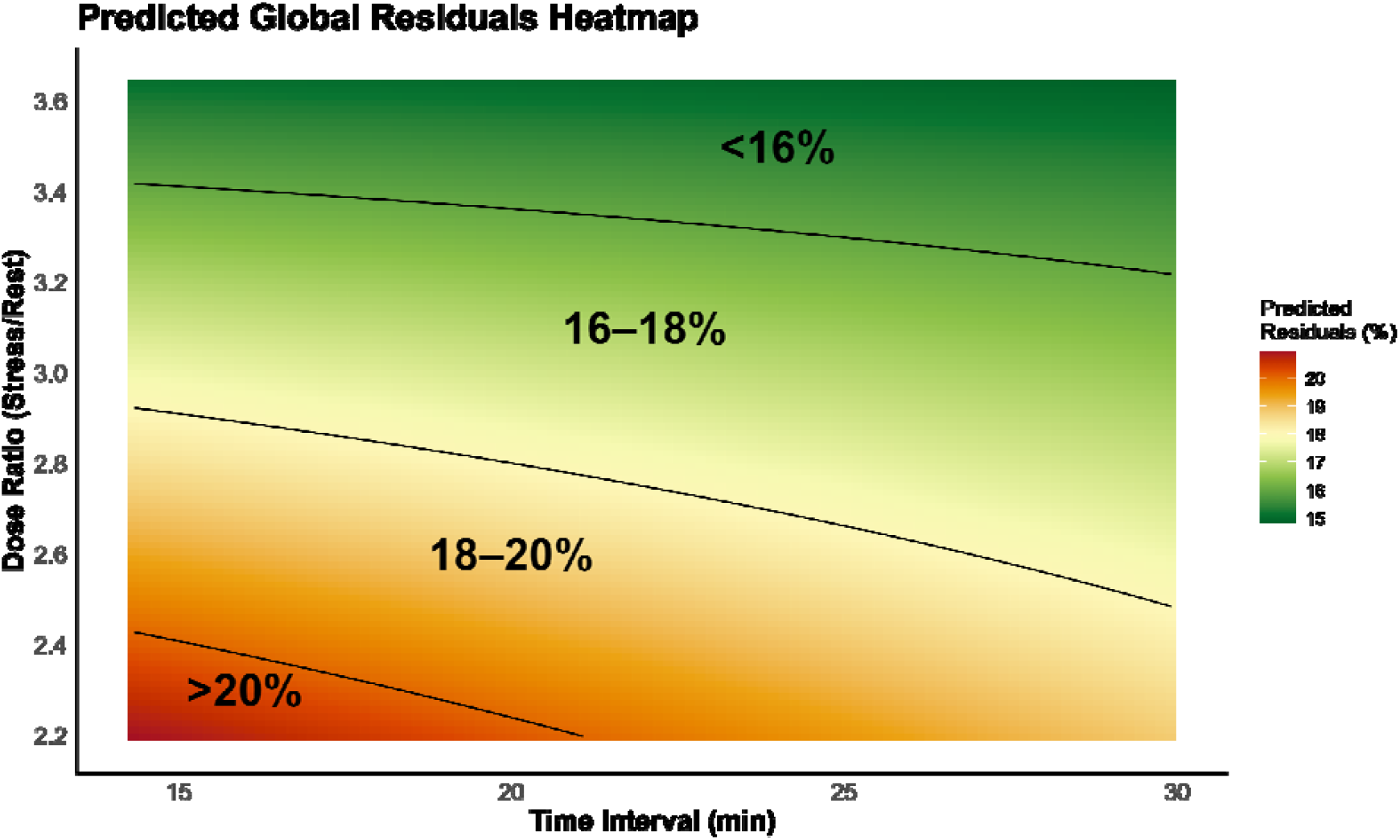
Predicted global relative residual activity (%) across varying stress-rest dose ratio and time intervals between injections.

### Optimal stress-to-rest dose ratio

The dose ratios at which the predicted probability of RRA<20% exceeded 90%,95% and 99% were 3.22, 3.55 and 4.28, respectively **(Figure 3)**. Linear regression cutoffs are shown in the Supplement **(Supplementary Figure 1)**. Global RRA was predicted at a fixed dose ratio of 3.2 across various time intervals, with all values consistently remaining below 20% **(Supplementary Figure 2)**.

**Figure 3.**
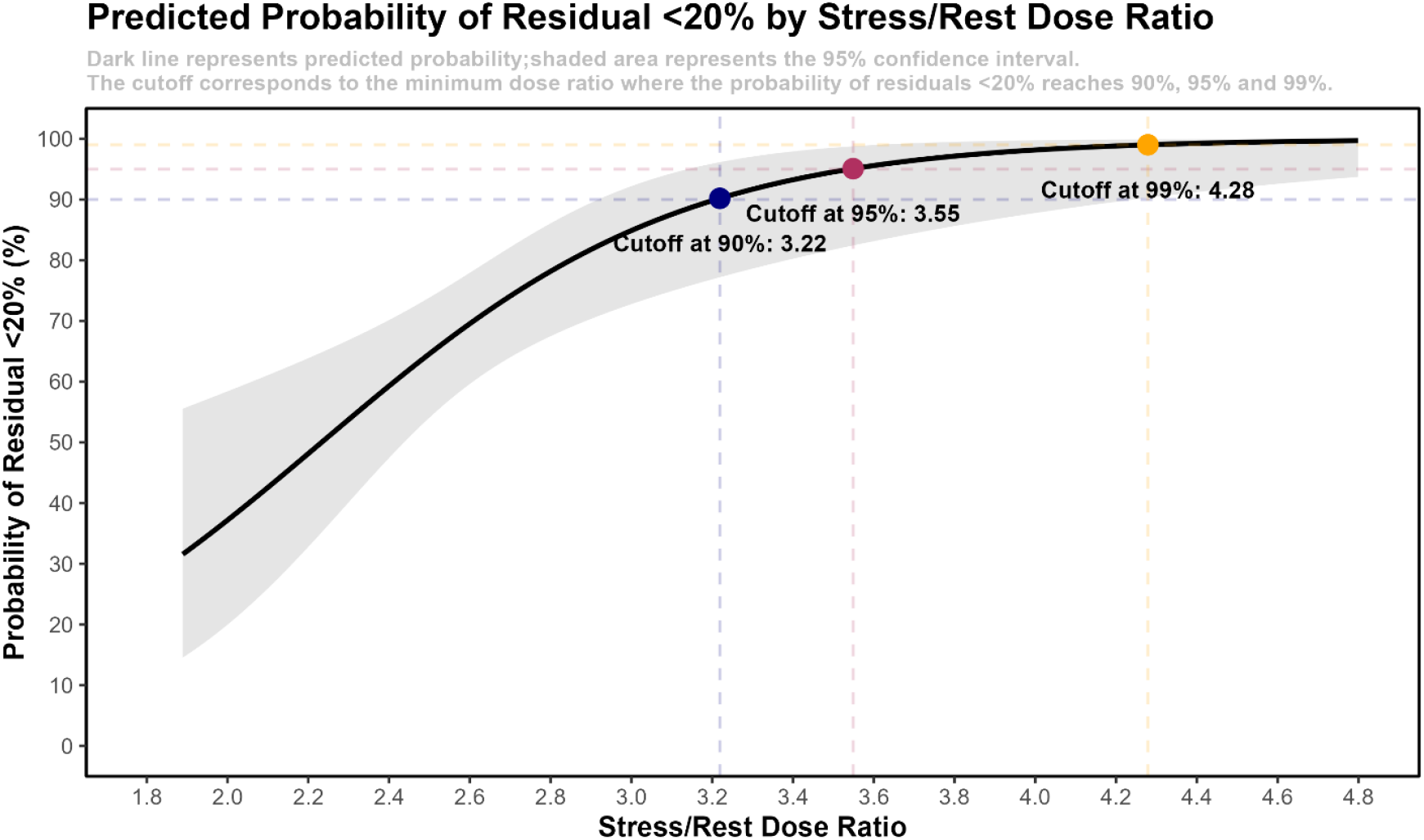
Predicted probability of global relative residual activity falling below 20% across dose ratios.

A mixed-effects model with a random intercept for each patient and an interaction term between dose ratio and vascular territory was used to assess whether the association between dose ratio and RRA varied across coronary territories. The analysis demonstrated that higher dose ratios were associated with lower residuals in all territories, with no significant interaction observed (P=0.9). The corresponding coefficients for each vascular territory are displayed in **Supplementary Table 1**.

### Visual assessment

The proposed back-to-back protocol, consisting of 10-minute rest and stress acquisitions for digital scanners with an optimized dose ratio and no intervening delay is illustrated in **Figure 4**. Representative images from this protocol are shown in **Figures 5A and 5B**, depicting patients with perfusion defects at dose ratios of 4.9 and 2.7, respectively. In both cases, the stress images demonstrate clear contrast despite the minimal delay between acquisitions.

**Figure 4.**
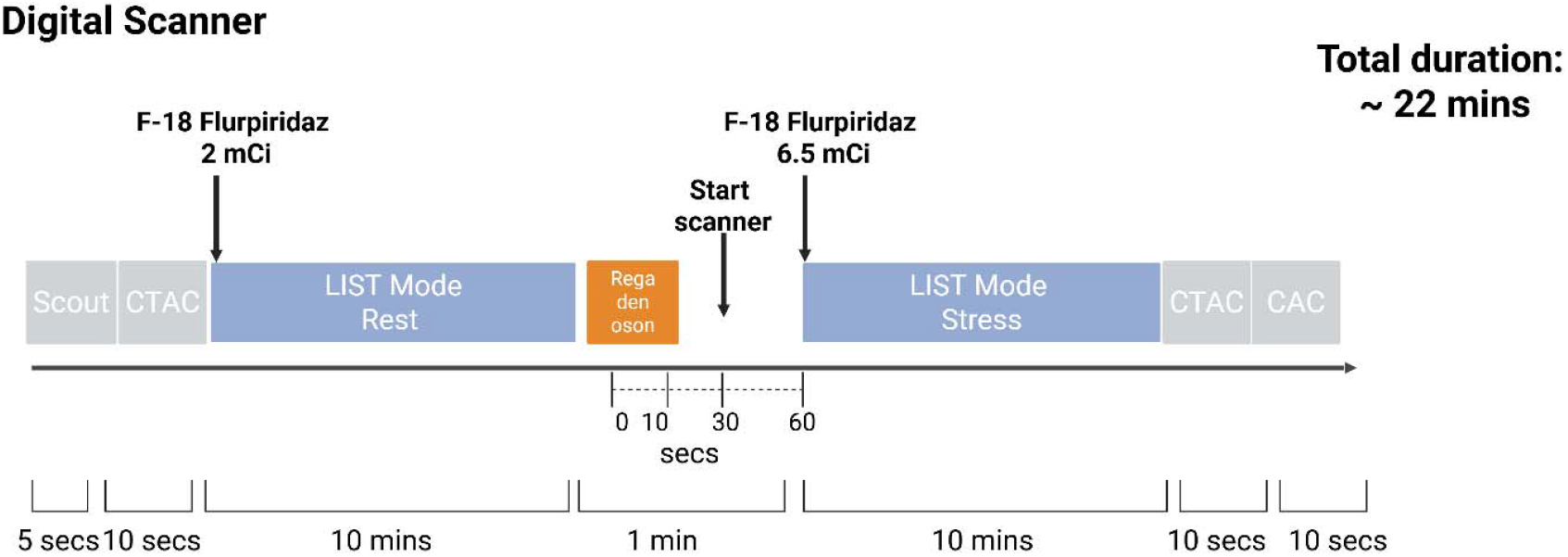
Suggested F-18 Flurpiridaz PET MPI protocol.

**Figure 5A.**
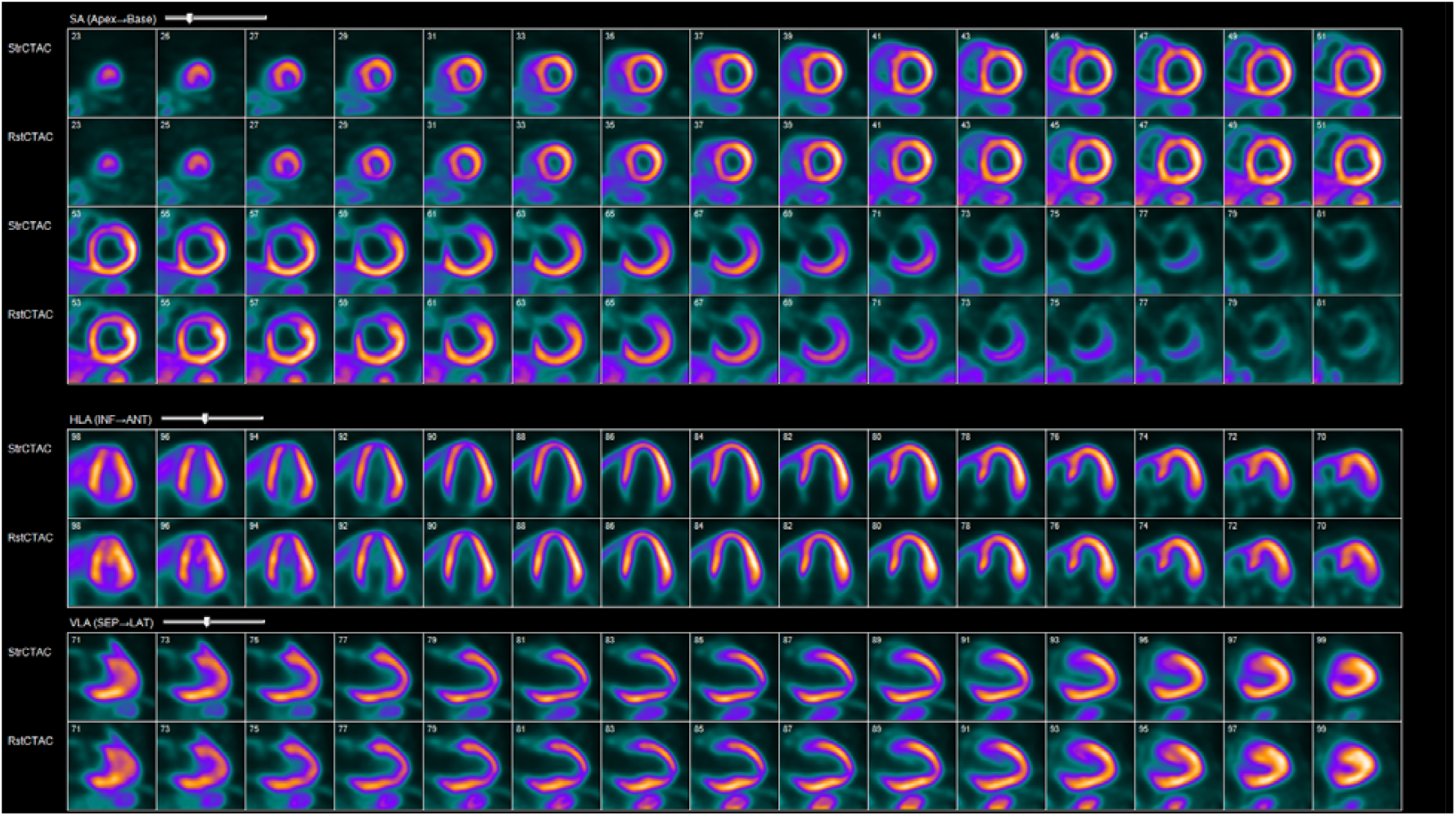
Study with 10 mins rest and stress acquisition, a wait time of 7 mins in between scans and a dose ratio of 4.9 in a patient with a BMI of 55 kg/m^2^.

**Figure 5B.**
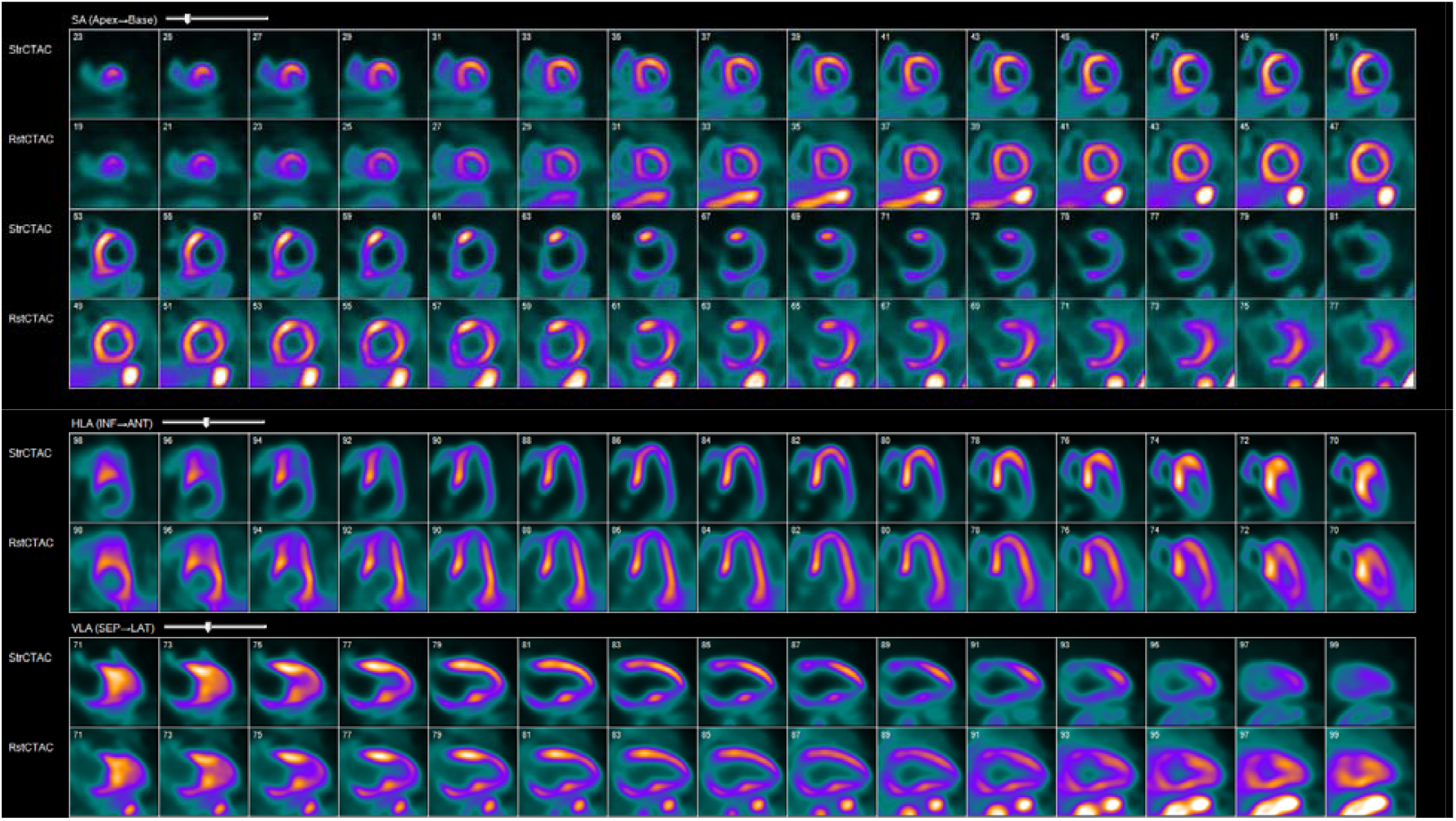
Study with 10 mins rest and stress acquisition, a wait time of 3 mins in between scans and a dose ratio of 2.7.

## Discussion

Our findings demonstrate that increasing the stress-to-rest dose ratio is strongly associated with reduced RRA, independent of the time interval between rest and stress injections. This relationship was consistent across all vascular territories. Further, the optimal stress-to-rest dose ratio required to ensure that residual counts fall below 20% ranged between 3.2 and 4.3.

F-18 has a relatively long half-life of 110 minutes, which is one of its key advantages, as it enables exercise stress testing and centralized unit-dose distribution.(2, 5, 6) These properties may support wider adoption of PET MPI, which, despite its proven efficacy, remains less widely adopted in the United States.(7, 8) However, this long half-life also extends the imaging protocol. To optimize image contrast between rest and stress and enable accurate detection of perfusion defects, it is essential to minimize the impact of the residual activity from the rest dose to ensure adequate visualization of the stress images. Prior protocols have recommended minimum delays of 30 minutes for pharmacologic stress and 60 minutes for exercise stress to allow residual rest activity to fall below 20% of the stress dose.(5, 6) However, these recommendations lack published validation and contribute to longer protocol durations for F-18 PET compared to Rubidium-82 (Rb-82) PET.

In a typical scenario, with a 2.5 mCi rest dose followed by a 6.0 mCi stress dose, a delay of ∼81 minutes between both injections would be required for the residual activity from the rest dose to fall below 20% of the total activity at the time of stress injection **(Supplementary material)**. This far exceeds the commonly used wait times of 30 minutes in clinical protocols, highlighting that prolonging the delay is not a practical solution in routine clinical workflows. Instead, increasing the dose ratio between the stress and rest doses emerges as a more practical strategy to enhance image contrast and allow for back-to-back imaging. Although dose ratios of approximately 1:2 for pharmacologic stress and 1:3 for exercise stress are often used in protocols,(6, 9) these values are based on empirical validation. Our findings provide data-driven support for dose ratio optimization, identifying a cutoff ranging between 3.2 and 4.3 to ensure that residual activity remains below clinically significant thresholds. While prolonging the wait time between injections could theoretically reduce RRA to <20%, our results show that stress-to-rest dose ratios between 3.2 and 4.3 achieve low residual activity regardless of the interval between injections. Thus, shorter wait times can improve workflow efficiency. For example, in an 8-hour day, a 10-minute delay (22-minute protocol for a digital scanner) would allow about 20 patients to be scanned compared with about 10 using a 30-minute delay (42-minute protocol). A longer protocol would be required for non-digital scanners, due to its lower sensitivity and time-of-flight as compared to digital scanners.(10)

Importantly, increasing the dose ratio by reducing the rest dose; rather than increasing the stress dose; offers the dual benefit of reducing radiation exposure for both patients and staff, while also shortening the overall protocol duration. Dose reduction is feasible because of the favorable physical properties of Flurpiridaz, including superior myocardial extraction, prolonged myocardial retention, and short positron range.(1)However, there is a limit to how much the rest dose can be reduced, which is highly dependent on scanner specifications. Dose ratio optimization allows for back-to-back imaging, thereby decreasing protocol time, increasing efficiency, improving patient experience, and enhancing co-registration of rest and stress data. Previous studies have explored alternative strategies to enable back-to-back imaging, including residual subtraction from stress images, with promising results.(11)

### Limitations

This study has several limitations. First, it was a single-center study using one type of scanner (digital). Second, the sample size was relatively small, especially in the group with longer time interval between rest and stress injections which may explain the non-significant association between wait time and RRA. Although longer delays could likely reduce RRA to <20%, this was not the focus of our study, which primarily aimed to show that RRA <20% can be achieved by optimizing the dose ratio alone regardless of the time interval between injections Third, the 20% threshold for acceptable RRA activity, is based on expert consensus rather than data and may warrant future validation based on diagnostic accuracy or image quality metrics. Fourth, we did not include patients who underwent exercise PET, and therefore, the proposed thresholds will require further validation in that setting.

## Conclusion

Our findings demonstrate that increasing the stress-to-rest dose ratio is a practical effective strategy to minimize the relative residual activity from the rest injection, consistently reducing it to below 20% regardless of the interval between injections. A dose ratio of 3.2 to 4.3 reliably enables back-to-back imaging without the need for prolonged delays, improving protocol efficiency and highlighting the need to streamline F-18 imaging protocols.

## Data Availability

All data produced in the present study are available upon reasonable request to the authors

## Abbreviations

PET: Positron Emission Tomography
MPI: Myocardial Perfusion Imaging
CT: Computed Tomography
F-18: F-18 Flurpiridaz
RRA: Relative Residual Activity

## New Knowledge Gained

### NEW KNOWLEDGE GAINED AND CLINICAL IMPLICATIONS

**What is new?**

- We demonstrate that increasing the stress-to-rest dose ratio reduced relative residual, independent of wait time.
- An optimal dose ratio of 3.2 to 4.3 consistently reduced relative residual activity to below 20%, enabling efficient back-to-back imaging.

**What are the clinical implications?**

- Dose ratio optimization offers a practical strategy to optimize F-18 PET MPI protocols by reducing protocol time and improving efficiency.
- As F-18 Flurpiridaz has only recently been FDA-approved, these findings may facilitate its wider clinical adoption by addressing workflow barriers.

## Graphical Abstract

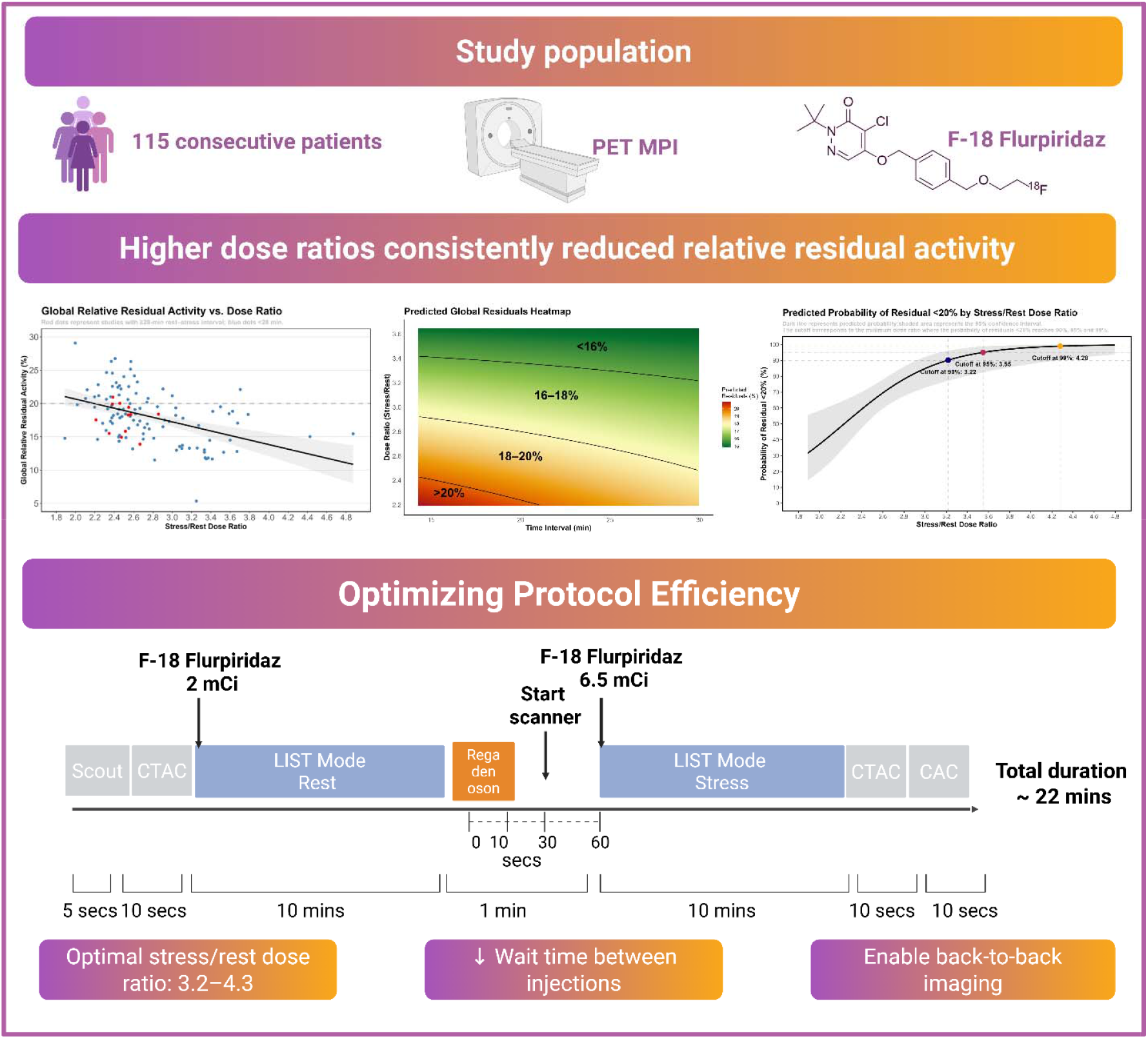

