## Supplementary Material for "Optimizing Protocol Efficiency in F-18 Flurpiridaz PET MPI Through Dose Ratio–Driven Reduction of Residual Activity"

**Author Line:** Maria Alwan, MD^1^; Ahmad El Yaman, MD^1^; Mahmoud Al Rifai, MD, MPH^1^; Sandra Escobar, CNMT, ARRT (N), (CT)^1^; Shah F. Abbasi, BS, ARRT (N), NMTCB^1^; Mohamad G. Ghosn, Ph.D.^2^; Marcelo F. Di Carli, MD^3^; Mouaz Al-Mallah, MD, MSc^1^

**Affiliations:**

^1^ Houston Methodist DeBakey Heart & Vascular Center, Houston, TX, USA

^2^ GE Healthcare

^3^ Brigham and Women's Hospital, Harvard Medical School, Boston, Massachusetts, USA.

**Address for correspondence:**

**Mouaz Al-Mallah, MD**
Houston Methodist Academic Institute, Weill Cornell Medicine,
Houston Methodist DeBakey Heart and Vascular Center
6550 Fannin Street, Smith Tower - Suite 1801,
Houston, TX, 77030, USA

**Disclosures:** Dr. Al-Mallah receives research support from Siemens and GE Healthcare. He is also a consultant to GE Healthcare and Jubilant.

**Ethical approval:** This study was approved by the Institutional Review Board at Houston Methodist Hospital.

**Funding:** None.

### Supplemental Methods & Results

To calculate the optimal stress–rest dose ratio, we first used cutoffs derived from logistic regression, identifying the ratio at which the predicted probability of relative residual activity fell below 20%. However, logistic regression treats all relative residual activity below 20% equivalently (e.g., 10% and 19% both count as “success”), without differentiating within that range. To further refine our analysis, we explored cutoffs based on the confidence interval (CI) and prediction interval (PI) from linear regression.

The 95% CI from linear regression reflects the mean residual for the population. The minimal dose ratio at which the upper bound of the 95% CI of the predicted relative residual activity fell below 20% was 2.43. However, this estimate represents the population mean rather than individual values; thus, while the mean is <20%, some individuals may still have residuals above this threshold. In contrast, the prediction interval reflects the range within which an individual’s residual is likely to fall, making it inherently wider. The minimal dose ratio at which the upper bound of the 95% prediction interval fell below 20% was 4.34, which was close to the cutoff obtained from the 99% logistic regression probability**. (Supplementary Figure 1)**

### Supplemental Figures

**Supplementary Figure 1. Predicted global relative residual activity (%) across dose ratios.**
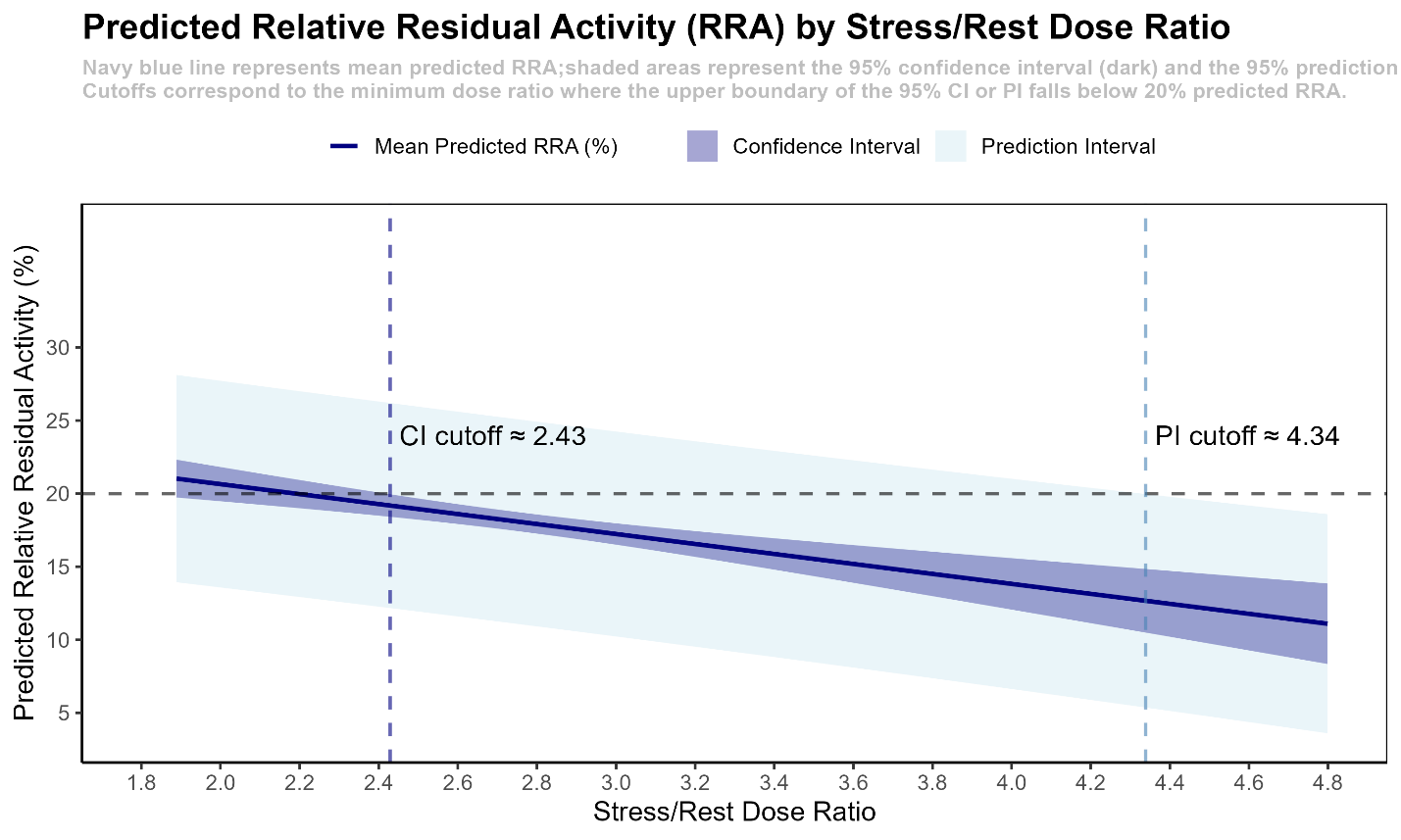

**Supplementary Figure 3. Predicted global relative residual activity (%) across time interval (mins) with a fixed dose ratio of 3.2.**

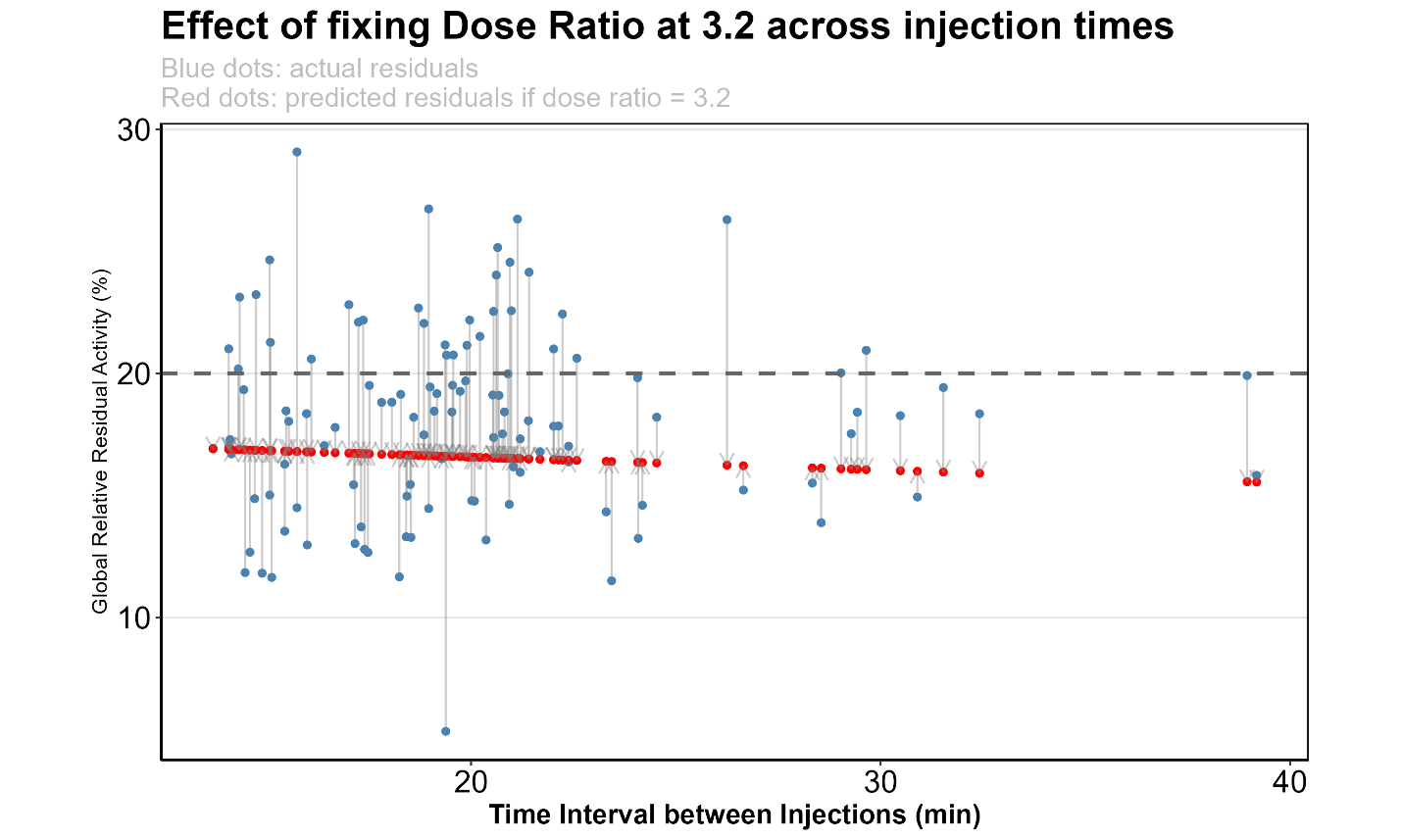

### Supplemental Table

**Supplementary Table 1. Association between dose ratio and relative residual activity across vascular territories. (Per 1-unit increase in dose ratio)**

| Territory | Dose ratio Coefficient (95% CI) | p-Value |
| --- | --- | --- |
| LAD | -3.35 (-4.88, -1.82) | <0.001 |
| LCX | -3.09 (-4.62, -1.56) | <0.001 |
| RCA | -3.32 (-4.85, -1.79) | <0.001 |

### Supplemental Discussion

**Calculation of wait time needed for rest dose to reach less than 20% of stress counts**

In a typical scenario, a rest dose of 2.5 mCi is injected followed by a stress dose of 6.0 mCi. To ensure that the residual activity from the rest injection contributes less than 20% of the total activity (stress dose + residuals), the condition can be expressed as:

   $\frac{R(t)}{S + R(t)}$ < 0.20

where R(t) is the residual activity at time t and S is the stress dose. Substituting R(t) with the standard radioactive decay equation:

R(t) = R_0_ × $e^{-\lambda t}$

where:

- R_0_ represents the initial rest dose at injection (2.5 mCi in this example)
- λ represents the decay constant and is calculated as λ = $\frac{ln(2)}{T½}$ ≈ 0.0063 per minute
- T½ represents the physical half-life of F-18 (≈110 minutes)
- t represents the elapsed time since the rest injection

Rearranging gives:

2.5 × $e^{-\lambda t}$
  ————————	< 0.20
  6.0 + 2.5 × $e^{-\lambda t}$

2.5 × $e^{-\lambda t}$< 1.2 + 0.5 × $e^{-\lambda t}$

2.0 × $e^{-\lambda t}$< 1.2

$e^{-\lambda t}$< 0.60

t = ln(1 / 0.60) / 0.0063 ≈ 81 minutes

Therefore, a delay of approximately 81 minutes would be required for the residual activity from the rest dose to contribute to just less than 20% of the total activity at the time of stress injection.
